# Opportunities for shared decision-making about major surgery: findings from a multi-method qualitative study of decision-making about orthopaedic, colorectal and cardio thoracic surgery with high risk patients

**DOI:** 10.1101/2022.08.02.22278194

**Authors:** Sara E Shaw, Gemma L Hughes, Rupert Pearse, Ester Avagliano, James R Day, Mark E Edsell, Jennifer A Edwards, Leslie Everest, Timothy J Stephens

## Abstract

**Background:** Little is known about the opportunities for shared decision-making when high-risk patients (over 60 years, with co-morbidities) are offered major surgery. This paper examines when and why clinicians and patients can share decision-making about major surgery.

**Methods:** Multi-method qualitative study, combining video-recordings of pre-operative consultations, interviews and focus groups (with a maximum variation sample of 31 patients, 19 relatives, 37 clinicians), with observations of clinics in five UK hospitals undertaking major joint, colorectal and/or cardiac surgery.

**Results:** Three opportunities for shared decision making about major surgery were identified. Resolution-focused consultations (cardiac/colorectal) resulted in a single agreed preferred option related to a potentially life-threatening problem, with limited opportunities for shared decision-making. Evaluative and deliberative consultations offered more opportunity. The former focused on assessing the likelihood of benefits of surgery for a presenting problem that was not a threat to life for the patient (e.g. orthopaedic consultations) and the later (largely colorectal) involving discussion of a range of options while also considering significant comorbidities and patient preferences. The extent to which opportunities for shared decision-making were available, and taken up by surgeons, was influenced by nature of the presenting problem, clinical pathway and patient trajectory.

**Conclusion and relevance:** Decisions about major surgery are not always shared between patients and doctors. The nature of the presenting problem, comorbidities, clinical pathways and patient trajectories all inform the type of consultation and opportunities for sharing decision-making. This has implications for clinicians, with shared decision-making about major surgery most feasible when the focus is on life-enhancing rather than life-saving treatment.

## INTRODUCTION

Shared decision-making is a collaborative process: clinicians and patients work together to share information about treatment and management options, consider preferred outcomes and reach agreement on the best care package for the patient.^1^ In the US federal legislation includes provisions for shared decision-making, with some states promoting it as essential to health care improvement.^3^ In the UK a landmark legal case in 2015^4^ expedited the shift to shared decision-making, focusing on what a patient would reasonably want or need to know.^5-7^ Guidance has followed,^8^ along with a parallel shift to patient-centred care.^10-12^ Other countries have followed suit.

Systematic reviews^13,14^ show that patients and clinicians value shared decision-making, patients tend to prefer it, and that it has potential to improve the quality of decisions (e.g. via information sharing) and reduce conflict around preference-sensitive treatment decisions about surgery (i.e. where there are two or more available options and no one best available treatment).

Little is currently known about shared decision-making for major surgery with high-risk patients. This is despite surgical treatments being increasingly offered to older patients and those with severe long term illness who are often at high risk of post-operative complications.^18,19^ Even when surgery and anaesthesia are straightforward, one in three high-risk patients develops serious medical complications (e.g. pneumonia) shortly after surgery,^20^ delaying recovery and extending hospital stays. Many high-risk patients never recover, suffering significant reductions in long-term quality of life and survival.^20,21^ Some experience regret over the decision to undergo surgery.^22^ Doctors want to improve decision-making for this patient group but are often ill-equipped to do so.^23^ Clinicians and patients are being asked to talk about decisions, yet they sometimes lack the knowledge and/or expertise to make informed judgements about how to balance longer term consequences with the need to address acute problems. There remains a dearth of literature assessing impact of acuity on decision-making processes, preferences and outcomes.^24^

There is a small but growing literature on the process of shared decision-making with high-risk patients.^25-30^ What literature there is suggests that high-risk patients often don’t realise they have a choice about surgery, and have unrealistic expectations about post-operative recovery. In this paper, we ask how, when and why do clinicians and high risk patients share decision-making about major surgery?

## METHODS

We used qualitative methods to explore the process of decision-making and activities allied to it. We have previously published a study protocol.^31^ This section provides a summary.

### Ethics and governance

The study received ethical approval from South Central Oxford C Research Ethics Committee (19/SC/0043) in February 2019, followed by governance approval from sites.

The study is part of the OSIRIS research programme (*Optimising Shared decision-makIng for high RIsk major Surgery*, https://osiris-programme.org/). The OSIRIS Steering Committee maintained oversight of the research. The OSIRIS Patient Panel informed study design.

### Setting

The research was conducted in five NHS hospitals (Table 1) undertaking two of three surgical procedures – major joint, colorectal, cardiac surgery – with diversity in location, population and hospital size.

**Table 1:**
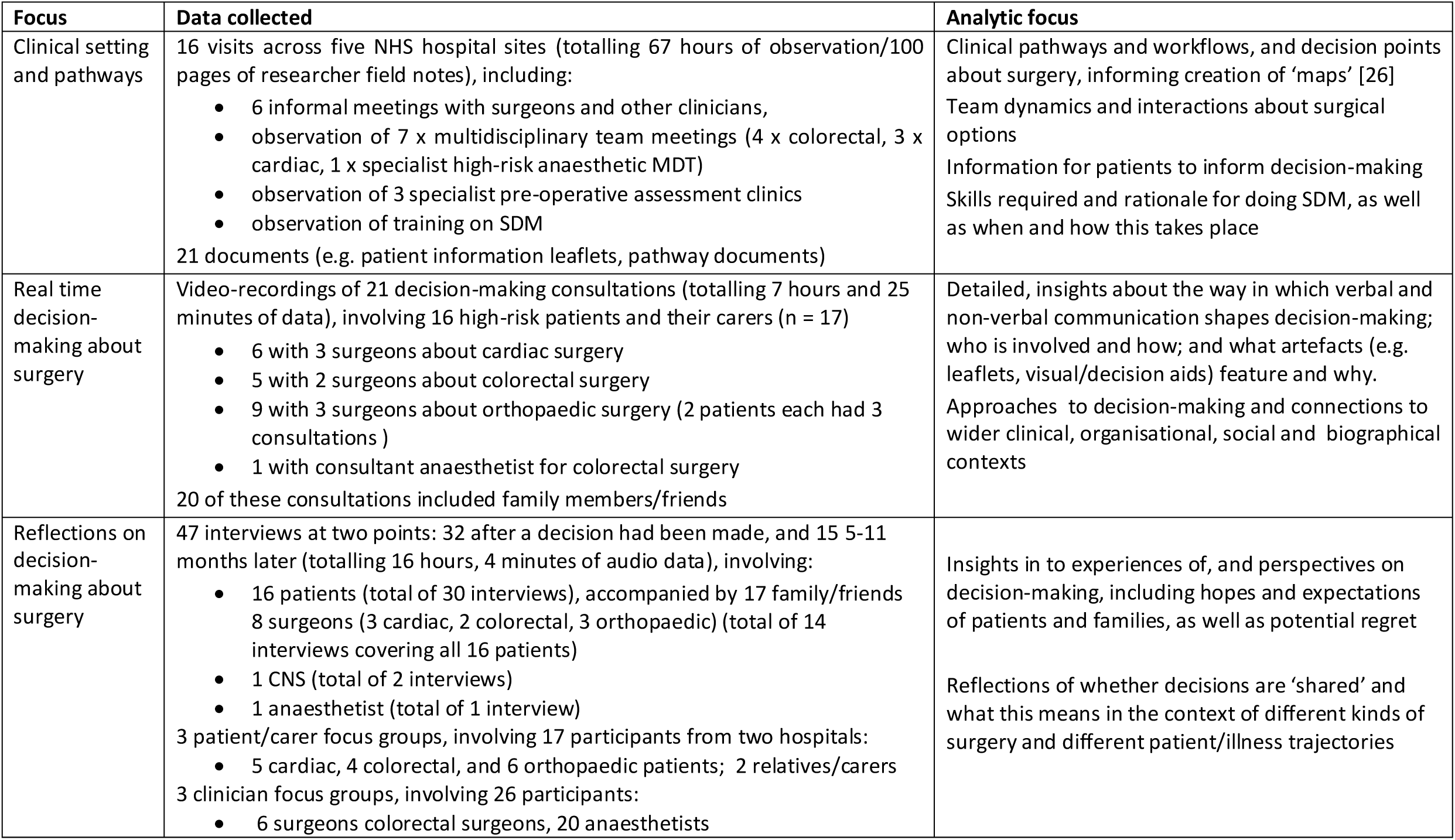
Overview of data collection and analysis.

| Focus | Data collected | Analytic focus |
| --- | --- | --- |
| Clinical setting and pathways | 16 visits across five NHS hospital sites (totalling 67 hours of observation/100 pages of researcher field notes), including:<br>• 6 informal meetings with surgeons and other clinicians,<br>• observation of 7 x multidisciplinary team meetings (4 x colorectal, 3 x cardiac, 1 x specialist high-risk anaesthetic MDT)<br>• observation of 3 specialist pre-operative assessment clinics<br>• observation of training on SDM<br>21 documents (e.g. patient information leaflets, pathway documents) | Clinical pathways and workflows, and decision points about surgery, informing creation of ‘maps’ [26]<br>Team dynamics and interactions about surgical options<br>Information for patients to inform decision-making<br>Skills required and rationale for doing SDM, as well as when and how this takes place |
| Real time decision-making about surgery | Video-recordings of 21 decision-making consultations (totalling 7 hours and 25 minutes of data), involving 16 high-risk patients and their carers (n = 17)<br>• 6 with 3 surgeons about cardiac surgery<br>• 5 with 2 surgeons about colorectal surgery<br>• 9 with 3 surgeons about orthopaedic surgery (2 patients each had 3 consultations )<br>• 1 with consultant anaesthetist for colorectal surgery<br>20 of these consultations included family members/friends | Detailed, insights about the way in which verbal and non-verbal communication shapes decision-making; who is involved and how; and what artefacts (e.g. leaflets, visual/decision aids) feature and why.<br>Approaches to decision-making and connections to wider clinical, organisational, social and biographical contexts |
| Reflections on decision-making about surgery | 47 interviews at two points: 32 after a decision had been made, and 15 5-11 months later (totalling 16 hours, 4 minutes of audio data), involving:<br>• 16 patients (total of 30 interviews), accompanied by 17 family/friends<br>• 8 surgeons (3 cardiac, 2 colorectal, 3 orthopaedic) (total of 14 interviews covering all 16 patients)<br>• 1 CNS (total of 2 interviews)<br>• 1 anaesthetist (total of 1 interview)<br>3 patient/carer focus groups, involving 17 participants from two hospitals:<br>• 5 cardiac, 4 colorectal, and 6 orthopaedic patients; 2 relatives/carers<br>3 clinician focus groups, involving 26 participants:<br>• 6 surgeons colorectal surgeons, 20 anaesthetists | Insights in to experiences of, and perspectives on decision-making, including hopes and expectations of patients and families, as well as potential regret<br>Reflections of whether decisions are ‘shared’ and what this means in the context of different kinds of surgery and different patient/illness trajectories |

### Sampling

#### Patients

We recruited 31 patients and 19 carers.

We first identified patients aged 60 or over who were considered high risk with a Charlson Co-morbidity Index (CCI)^32^ score ≥4 indicating significant co-morbidities or clinically frail (for cardiothoracic surgery, patients’ risk was primarily related to their cardiac problem - Table 2). We recruited 16 patients (5 orthopaedic, 5 colorectal, 6 cardiac), across 3 sites who were currently undergoing care, ensuring diversity in age, gender and social circumstances. Fifteen patients were accompanied by at least one carer (n =17).

**Table 2:** Summary of patient characteristics, conditions and decisions.

| Patient age range | M/F | CCI | Presenting problem | Consultations observed | Accomp'd by | Decision made | Type of decision-making |
| --- | --- | --- | --- | --- | --- | --- | --- |
| **Colorectal patients (n=5)** |  |  |  |  |  |  |  |
| 76-80 | M | 5 | Positive result from bowel cancer screening | 1, with consultant surgeon + CNS | Wife | Major surgery (right hemicolectomy to remove tumour and rejoin bowel) | Resolution-focused |
| 71-75 | M | 5 | Anaemic | 1, with consultant surgeon + CNS | Wife | Major surgery (right hemicolectomy to remove tumour and rejoin bowel) | Resolution-focused |
| 76-80 | M | 5 | Anaemic, history of stomach cancer | 1, with consultant surgeon | Wife + sister | Transanal minimally invasive surgery to remove part of rectal tumour | Deliberative |
| 76-80 | M | 8 | Abnormal colon detected during scan for respiratory problems | 1, with consultant surgeon | Wife | Surveillance (colonscopy) | Deliberative – shifting to evaluative |
| 61-65 | F | 6 | Weight loss, escalating rapidly to pain (previously not disclosed to clinical team) | 2, with consultant surgeon and consultant anaesthetist | Daughter | Emergency surgery to address obstructing bowel and create stoma | Deliberative |
| **Orthopaedic patients (n=5)** |  |  |  |  |  |  |  |
| 76-80 | F | 5 | Pain and mobility problems in hips (bilateral hip replacement 21 years previously) | 3, with consultant surgeon | Son | Watch and wait follow-up appointments with consultation surgeon | Evaluative |
| 71-75 | M | 4 | Knee pain (replacement knee 13 years previously) | 3, with consultant surgeon | Wife | Surgery to resurface kneecap –with option to revise joint if required | Evaluative |
| 66-70 | F | 3+ | Knee pain (replacement knee 5 years previously) | 1, with consultant surgeon | No | Further investigations / watch and wait | Evaluative |
| 66-70 | M | 4+ | Knee pain (replacement knee 18 months previously) | 1, with consultant surgeon | Wife | Further investigations / watch and wait | Evaluative |
| 81-85 | F | 5 | Knee pain | 1, with consultant surgeon | Friend | Knee replacement | Evaluative |
| **Cardiac patients (n=6)** |  |  |  |  |  |  |  |
| 71-75 | F | 3 | Breathlessness and fatigue | 1, with consultant surgeon | Husband | Aortic valve replacement | Resolution focused |
| 66-70 | M | 3 | Chest pain | 1, with consultant surgeon | Wife | Bypass surgery | Resolution focused |
| 76-80 | M | 3+ | Aortic aneurysm detected during scan for spinal problems | 1, with consultant surgeon | Wife + daughter | Surgery not advised | Resolution focused |
| 71-75 | M | 3 | GP detected 'murmur' and referred to cardiologist | 1, with consultant surgeon | Wife | Mitral valve repair | Resolution focused |
| 81-85 | F | 5 | Aortic aneurysm detected during investigations for breathlessness, referral to cardiac surgeon had been delayed as multiple myeloma diagnosed in meantime | 1, with consultant surgeon | Husband | Updated scans required, investigations ongoing | Evaluative (because of lack of info) |
| 66-70 | M | 3+ | Under care of cardiologist- has heart disease and PCI previously | 1, with consultant surgeon | Wife | Referral to respiratory specialist (now lost to follow-up) | Resolution-focused *but* ended with evaluative as referred on |

We recruited a further 17 participants for three patient / carer focus groups about past experiences of decision-making about surgery and 26 participants for three anesthetist /surgeon focus groups (Table 1). Clinicians were invited to focus groups via Royal Colleges and professional networks. Written informed consent was obtained from all participants.

### Data collection

We video-recorded 21 consultations (10-45 minutes) that involved decision-making about major surgery with 16 patients and their carers (Jun-19 to Jan-20). Two colorectal patients were seen at a hot clinic (a ward-based service for those needing urgent treatment), with 14 seen at outpatient clinics.

The colorectal consultations (n=5) were planned to discuss options after investigations. Orthopaedic consultations (n=5), were part of on-going evaluations. For two patients we recorded three consultations over 5 months. Cardiac patients (n=6) had typically had non-surgical options ruled out.

Video-recording involved placing a camera in the consultation room to record interaction between the patient, anyone accompanying them (Table 2) and the clinician(s).

We conducted narrative interviews^33^ with clinicians and patients (plus carer where relevant) after each consultation, and 5-11 months later (i.e. after having/declining surgery). By the end of the study, due to COVID-19, one patient was still waiting for surgery, one patient had moved abroad. We held 6 focus groups (detail in Table 1).

### Analysis

As usual in qualitative research, analysis was informed by literature on shared decision-making^1,29^ and the social science of decision-making, acknowledging that decisions about surgery rarely occur at neat ‘decision points’,^35^ and involve deliberation^36,37^ and interaction^38-40^ at various points in the patient trajectory.

We mapped clinical pathways and decision-making processes,^31^ and combined this with video and interview data to produce case summaries. We then used thematic analysis^41^ and constant comparison^42^ to identify different types of consultation for major surgery and approaches to shared decision-making. Finally we examined the interactional order^39,43^ of all consultations: (i) mapping activities during each consultation, (ii) identifying the substance, form and rules^39^ for each consultation type, and (iii) conducting detailed analysis where options were discussed and decisions made.) mapped activities, examined the substance, form and rules^39^, and conducted detailed analysis where options were discussed and decisions made. We tested emerging analysis in focus groups.

## RESULTS

A significant amount of work took place before patients met with their surgeon to discuss treatment options. This was guided by clinical pathways (e.g. time to reach the surgeon),^31^ patients’ candidacy (i.e. eligibility for surgical attention/intervention^44^) and multi-disciplinary review.

All consultations with the surgeon included discussion about the nature of the problem, causation and prognosis and how it was affecting the patient, explanation of proposed surgery, immediate operative risks (e.g. infection), and what would happen after the consultation and after surgery. Beyond this consultations varied, falling into one of three types - resolution-focused, evaluative and deliberative– - each with different opportunities for shared decision-making (Figure 1).

**Figure 1:**
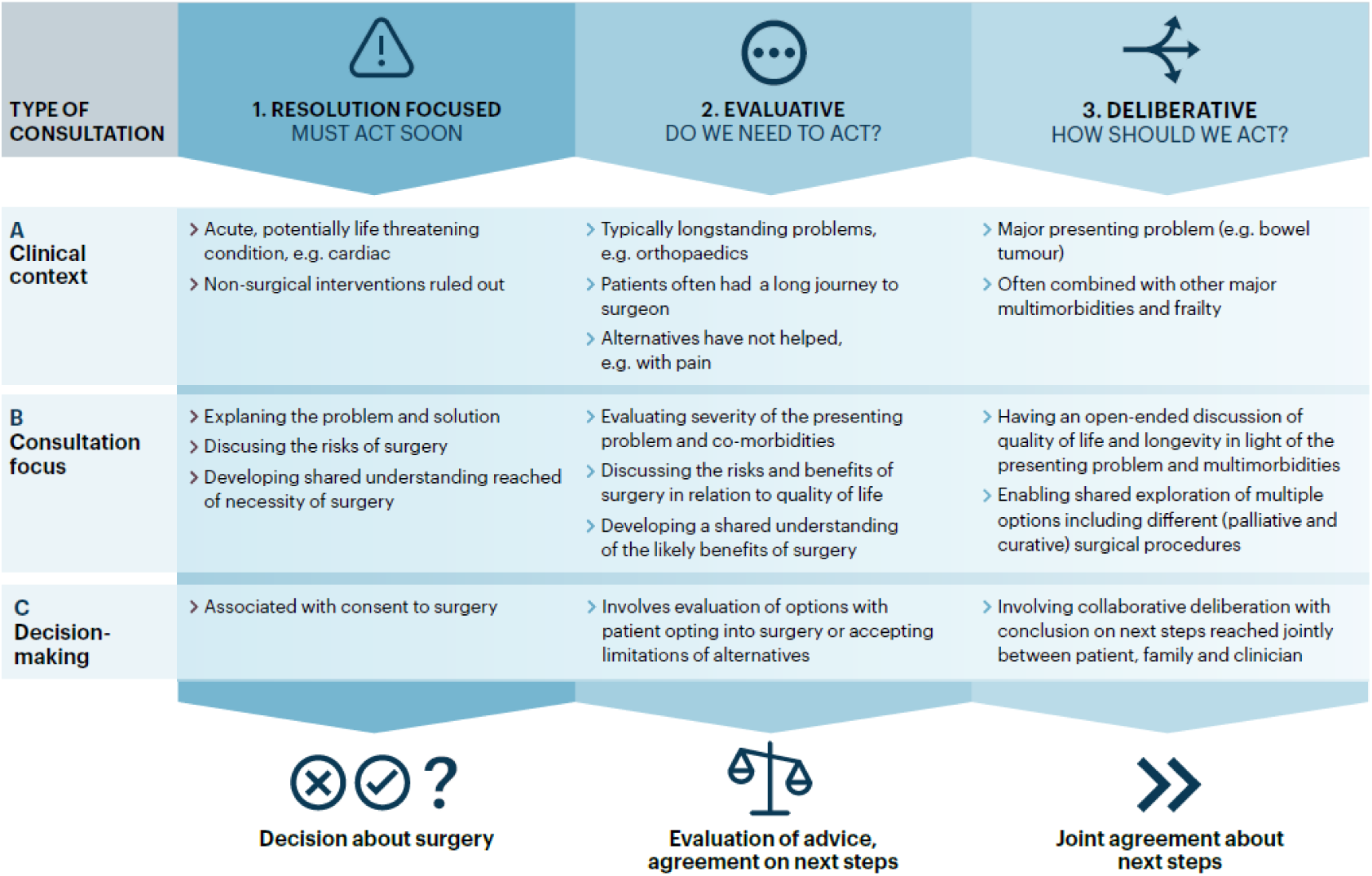
Opportunities for share decision-making about major surgery.

### Resolution-focused consultations

These consultations (5 cardiac problems, 2 bowel tumours – Table 2) typically took the form of a traditional consultation, reviewing medical history and explaining the problem/pathology ahead of discussion about treatment. Patients and clinicians both understood the problem to be potentially life-threatening, with surgical intervention the optimal route to resolving (or ‘fixing’)^45^ the problem and maximising the patient’s chances of survival (see Table 3 for exemplary quotes). In two cases, operative risks led the surgeon to recommend against surgery or refer for further investigations. Surgeons informed remaining patients about risks and reassured that surgery was major but routine.

**Table 3:** Data examples by consultation type, drawn from patient and clinician interviews and video-recordings of consultations.

|  | Resolution-focused consultations | Evaluative consultation | Deliberative consultation |
| --- | --- | --- | --- |
| Presenting problem | *I had an annual check-up, where they did a blood test. And it was discovered that I was very anaemic...which automatically triggers endoscopy and then they found the polyp*<br>P02, interview<br>*I feel great! But I get out of breath... so breathless and I get tired... and when I get short of breath and it tightens all up in my chest*<br>P10 to cardiac surgeon | *I'm having problems getting up the stairs...there's always been pain*<br>P03 interview<br>*I can definitely feel it starting from sort of here coming down...particularly if I am going downhill or uphill or upstairs or downstairs*<br>P04, consultation with orthopaedic surgeon<br>*...with the pain I'm getting... It stops me in my tracks if I'm doing something*<br>P07, interview | *I went to my doctor... and he put me on iron tablets because... and he said, 'I'm going to send you for a scope both up and down... and that's where it started. He said, 'They've obviously spotted something*<br>P05 during interview<br>*pain...vomit...we've realised ...the gut is quite different from when it was when we saw you*<br>surgeon to P14 |
| Clinician framing of the problem | *this is a very common cancer...most people with it are cured... the treatment...that we'd recommend would be an operation*<br>Colorectal surgeon to P01<br>*if we try to operate on this one your chance of stroke will be very high*<br>Cardiac surgeon, consultation with P13<br>*I didn't think of it as a decision to be made. When they tell you you've got cancer and we can operate to remove it...that was no decision*<br>Patient with colorectal cancer, Focus Group 6 | *your knee is now not functioning quite so well but it's ok... you are functioning pretty well*<br>Orthopaedic surgeon, consultation with P07<br>*it's never an operation that you have to have done it's not life-saving. It's meant to improve things for you...but there is a possibility that it could make things worse*<br>Orthopaedic surgeon, consultation with P09 | *He tried to convince me last time that he was really fit and well... there was some warning bells...An early rectal cancer...opens up the option...we were talking about*<br>Colorectal surgeon (about P05), interview<br>*So, I start with a similar sentence... 'Do you know why you've come to see me?' and I just stay quiet, and most of them go, 'Well I'm not really very fit, am I?' To begin with that pause, they'll fill it in and you're on the right page with them...*<br>Anaesthetist, focus Group 1 |
| Patient understanding | *Once I knew that I had the cancer, then it was a matter of coming to terms with that, and getting it sorted as quickly as possible... now it's a matter of dealing with what's there*<br>P01, interview | *I can't explain how excruciating it is...but it's only for a short period... ..but I mean if it doesn't get any worse I can tolerate that*<br>P07, to orthopaedic surgeon | *Well I was told there were 3 options.*<br>P05, consultation with colorectal surgeon<br>*I didn't know they were going to cut as much away*<br>P06, interview |
| Discussing choices | *we obviously always need to consider the other options but...there's not really any other* | *'... so that's the plan to re-surface the knee cap and or revise the whole knee that's one* | *Do you want the major surgery...or the [local surgery]? It's not something that's offered to* |
|  | *surgical options...there is the option of doing nothing*<br>Colorectal surgeon, consultation with P01<br>*I mean there's an issue here about when I get in a taxi and he asks me which route do you want me to take, and I say, 'Well hang on a minute, you're the taxi driver'*<br>Focus Group 1, Cardiac patient | *side...the second one is wait and watch...there's two approaches to this...'*<br>Orthopaedic surgeon consultation with P04<br>*it's your decision...I'll help you try to get to what you feel is the right thing ...but it's there is quite a lot of uncertainty so you have to have a bigger operation with a longer recovery where there's a risk of...complication blood clot, infection, problems with your heart and your lungs these sort of things.*<br>Orthopaedic surgeon, consultation with P08 | *everyone but your results so far are going to suggest that's an option*<br>Colorectal surgeon, consultation with P05<br>*if we chose nothing [we'd]...have a chat with the palliative care doctors and see if we manage that just as comfortably as we can for you...but it's not going to treat it and it's not going to take anything away... you've got a couple of surgical options so they all involve a general anaesthetic*<br>Colorectal surgeon, consultation with P14 |
| Articulating risk | *It's a fairly big operation but we do it routinely and I'll make sure you do well... the overall risk of the operation is about 2% so success is 98%*<br>Colorectal surgeon, consultation with P10<br>*there are risks involved... the risk of surgery is probably about 1% and...stroke I think you have a 98 99% chance of coming through the operation ....less serious problems ... there's a risk of infection...chest, urine infection*<br>Cardiac surgeon, consultation with P15 | *So the main risks that you need to know about are infection...so that's about 1% probably slightly higher because you're on the warfarin... despite the fact that you'll be on your warfarin, there is still that risk of blood clots...the other one the other big one is ongoing pain and stiffness so it might not give you the result that you want from it...there are some other ones that are more minor*<br>Orthopaedic surgeon, consultation with P09 | *It's explaining what risk is. You can say someone's high risk but you have to say high risk and all the complications; higher risk of a longer hospital stay, higher risk of not getting back to your current function capacity, needing carers.*<br>Focus Group 1, anaesthetist |
| Reaching a decision | *in my case he can't do anything... you've just got to face the facts*<br>P13, interview<br>*really I've got no choice...I want a better quality of life...although I don't like it...I've got a problem...get it fixed*<br>P15, consultation with cardiac surgeon<br>*It was easy. The pain decided for me*<br>Focus group 1, orthopaedic patient | *I wouldn't mind going for the [surgery]*<br>P04, consultation with orthopaedic surgeon<br>*I was hoping it wouldn't come to that but at the same time if its needs done and its going give me less pain and a bit more mobility I would do it*<br>P09, consultation with orthopaedic surgeon | *I personally feel that's a better option if you could handle it, but it's you that's got to go for the decision. [yes] I'll go for that then*<br>P06 wife and P06 during consultation<br>*you have ruled out the 'no' I think haven't you? And you're kind of ruling out, I think, the more extreme longer version...you're in the middle*<br>Colorectal surgeon, consultation with P14 |

These consultations focused on discussing a preferred option, typically surgery. Surgeons had a clear view of the presenting problem and potential benefits of surgery, informed by diagnostics and multi-disciplinary review. Other procedures (e.g. PCI for cardiac patients) had already been ruled out. Patients came to the consultation with an understanding that they had a serious condition that needed fixing. While patients were offered a list of options, (including ‘doing nothing’), there was general agreement that surgery was the optimum choice if the patient was fit enough. The content of resolution-focused consultations focused largely on medical knowledge, appreciation of pathology and weighing up of operative risks. Across consultations there was significant discussion of the problem, with artefacts (e.g. scans) used to aid explanation. From the surgeon’s perspective the aim was to help patients understand that surgery was being offered (in one case, not offered), maximise chances of survival and ensure patients were fully informed/able to consent to the proposed recommendation. Patients in focus groups unanimously agreed that this is what they experienced and expected.

In sum, the opportunity for shared decision-making in resolution-focused consultations centred around informing the patient about potential surgery and supporting them to make a decision about whether or not to accept it.

### Evaluative consultations

Evaluative consultations involved orthopaedic patients with persistent co-morbidities (n=5), plus one colorectal and one cardiac patient. Before coming to the surgeon orthopaedic patients had already consulted one or more health professionals and had non-surgical interventions (e.g. physiotherapy). Four presented with pain related to a previously replaced joint. All were hoping for resolution. One colorectal and one cardiac patient fell into this category due to information about comorbidities. Unlike resolution-focused consultations, there was no predetermined solution. The focus was on evaluating the patients’ situation and assessing options and benefits (Table 3).

From the surgeon’s perspective these were consultations about *life-enhancing*, rather than *life-saving*, treatments. Surgeons focused on evaluating if surgery was likely to help, and whether frail and multi-morbid patients would be worse off as a result. Decision-making focused on what was best for the patient. In all 5 orthopaedic consultations the surgeon’s knowledge guided encounters: surgeons typically clarified pathology and the likelihood of surgery helping, summarised the extent of problems experienced and associated pain, weighed up potential risks and benefits and cautioned about the likelihood of success of surgery. Potential risks and outcomes for treatment options were frequently (but not always – Table 3) quantified. Examination, plus imaging and models, was used by surgeons to assess and explain aetiology of the presenting problem.

In three consultations, the surgery was not an option. Patients were offered follow-up appointments and advice about managing their problem. Two orthopaedic patients were offered further investigations (scans, joint injections) and the colorectal patient further surveillance (endoscopy). These were framed as ‘watch and wait’ decisions, keeping the option for surgery open. With their clinicians, two patients reached an assessment that surgery could be of benefit and then confirmed that decision. The cardiac patient was referred for diagnostic tests.

In sum, opportunities for shared decision making in evaluative consultations involved developing a shared understanding of the benefits (or otherwise) of surgery, led by the clinician and often involving discussions over several consultations. Potential benefits/risks (in terms of surgery and patients’ quality of life) were evaluated relative to comorbidities. Patients were supported to make a decision, either accepting surgery or continuing with non-surgical management.

### Deliberative consultations

In deliberative consultations it was the high risk status of the patient combined with their presenting problem – in this study, bowel tumours – that was paramount when considering next steps (Table 3). Discussion about the potential benefits of surgery was explicitly linked with patients’ frailty and likely consequences (e.g. hospitalisation), with alternative surgical and non-surgical options considered alongside discussion about anaesthetic procedures. This opened up more opportunities for collaborative construction of options, and then shared decision-making, than in other types of consultation.

Co-construction of options was shaped through discussion of each patient’s situation, including co-morbidities and preferences, with collaborative deliberation^31^ of options focused on what survival might mean in the context of patients’ clinical, social and family situation (e.g. whether they may need long term carer support post-operatively). One surgeon led deliberative evaluations with 3 patients, all of whom had tumours for which surgery would usually be warranted but in these cases was thrown into question given comorbidities. Anaesthetists and colorectal surgeons in focus groups stressed the value of this approach for higher risk patients while acknowledging the challenges in doing this, in particular the time taken for such discussions.

Consultations were framed in terms of what mattered to patients. Longer term risks of harm, reduced quality of life and level of uncertainty were explicitly discussed, allowing patients, relatives and clinicians to weigh up risks, benefits and uncertainties. Operative risk was raised, including risk of mortality, likelihood of needing long term care and potential to live independently post-surgery.

Options were explored collaboratively. Patients were asked to make their decision from a list of options. Two patients opted for lesser (one palliative, one less invasive) surgical interventions. One consultation shifted into an evaluative discussion about how much the tumour was a problem in light of a respiratory problem, leading to a decision to ‘watch and wait’.

In sum, these consultations provided significant opportunity for shared decision making, with collaborative deliberation about risks, uncertainties and potential benefits integral. The selection of an option was down to the patient.

## DISCUSSION

### Summary of main findings

Guidance on shared decision-making suggests that every treatment decision has the potential to be shared. This study has shown that this is not the case for major surgery with patients at high risk of poor long term outcomes. The combination of qualitative methods, an explicit focus on interaction in consultations and sensitivity to the processes of decision-making involving high risk patients allowed us to: (i) reveal how colorectal, cardiac and orthopaedic surgeons adopt distinct and varied approaches to consulting with patients about major surgery; (ii) identify three types of consultation that offer different opportunities for shared decision-making, (iii) raise the possibility that shared decision making may not always be possible; and (iv) highlight that decisions may unfold over time and across multiple encounters.

### How findings add to the existing literature

Findings add to the small but growing literature on shared decision-making with high risk patients. This indicates that discussions between surgeons and patients about potential post-operative complications often have significant communication gaps,^20,21^ with reliance on surgical expertise and experience (i.e. over individual, preference-sensitive choice). Both parties tend to *assume* shared values, which shapes decision-making with, for instance, patients citing lack of belief in the surgeon’s prognosis as informing their decision.^22^ Our findings show that options in resolution-focused (or ‘fix it’^45^) consultations are perceived to be extremely limited (e.g. surgery or death) or non-existent. This does not mean that (for this group of high risk patients at least) resolution-focused consultations are not patient centred, but that they focus more on creating a shared understanding of surgery. Patients in these consultations wanted to have their problem fixed, saw that as the surgeons’ role and (whether they had surgery or not) were happy with the decision-making process and the decision made. As reported elsewhere, care is needed to avoid focusing on ‘fixing’ a problem in ways that close down discussions about the value of surgery;^45^ however, our findings suggest that shared decision making is not necessarily possible or desirable in resolution-focused consultations.

There is a risk that resolution-focused consultations play out this way due to ‘clinical momentum’^1,23,36^ and that surgeons make judgements about patients’ disposition for shared decision-making and act accordingly.^20^ Previous studies suggest a mis-match between clinician and patient preferences for participation in decision-making^24,37,38^ and between what surgeons discuss and what patients want to know (typically less technical information, and more on survival and longer-term quality of life)^39^. In such cases a more evaluative or deliberative approach may be appropriate.

To our knowledge, the delineation between different types of consultation for major surgery, the interaction involved and differential potential for shared decision making with high risk patients is new. In evaluative and deliberative consultations the focus was on life-enhancing treatment (albeit with high risk of poor longer term outcomes) with more opportunity, not only for identifying options, but also for discussing these in the context of each patient’s situation and preferences. This resonates with literature on collaborative deliberation.^31,40^

To date limited attention has been given to how shared decision making is shaped by family and other social factors.^37^ We did not set out to explore the specific role of family, however findings show that relatives were involved in shared decision-making, particularly in evaluative and deliberative consultations.

### Strengths and limitations

This is the first published study capturing qualitative evidence about interaction in decision-making consultations and the wider context in which that takes place. As is frequently the case in qualitative research, our patient sample was small. We used multiple methods to generate a rich dataset enabling in-depth analysis of consultations. Interactional data in particular, combined with interviews, has enabled detailed insights on the process of decision-making, allowing us to identify when and how decisions were made and the extent to which they were shared. Testing emerging analysis with a wider group of clinicians and patients in focus groups was helpful, albeit limited in terms of clinical speciality and patient experience. We used the Charlson Comorbidity Index^32^ to help identify high-risk patients. Some patients with lower CCI scores were considered by clinicians to be high-risk and vice versa. We sought to address this by working with recruiting clinicians to include patients identified as high-risk (frail). It is possible that the same study conducted in different sites would identify different kinds of ‘high risk’ patients.

### Conclusion

The dominant assumption is that shared decision-making is relevant to every consultation. Findings indicate that the traditional medical consultation is reinforced in resolution-focused consultations with limited focus on shared decision-making (which may be appropriate for some patients). Evaluative and deliberative consultations appear to provide greater opportunities for shared evaluation of the potential benefits of surgery in specific types of consultation. Deliberative consultations in particular are likely to be appropriate for older, frail patients for whom the longer term outcomes of surgery are uncertain. Surgeons are likely, at least implicitly, to be aware of the different types of consultation we found when they are consulting with high risk patients, differential opportunities for shared decision-making, and the challenges inherent in a more deliberative process.

Further research is needed to explore the extent to which the resolution-focused, evaluative and deliberative consultations are used, the opportunities created for shared decision making, and clinicians and patients perceptions about, and experiences of, how and when to use them. Different specialities will undoubtedly lend themselves to different types of consultation. Making the type of consultation explicit could help in appropriately enabling and supporting, if not always sharing, decisions.

## Data Availability

All data produced in the present study are available upon reasonable request to the authors

## DETAILS OF AUTHOR’S CONTRIBUTIONS

SS conceived the original study design, with input from RP, TS and LE (part of the wider Osiris programme, on which RP is Chief Investigator). SS is Principle Investigator for the study and guarantor for the article. EA, JD, ME and JE led on participant recruitment across study sites. GH collected data for interviews and video-recording of consultations, TS for focus groups, with input from SS and GH. SS and GH led on analysis and interpretation of data. TS analysed focus group data. SS drafted the article, and worked closely with GH, TS and RP to revise it. All authors commented on a draft version of this paper, approved the final version and agreed to be accountable for all aspects of the work.

## ACKNOWLEDGEMENTS

Our thanks go to all those involved in shaping and conducting the OSIRIS programme. Particular thanks go to all those who participated in this study, including clinicians, patients and their families without whom the research would not have been possible. We thank Lucas Seuren for advice and support on analysing communication and interaction; and John Prowle for his contribution to shaping this part of the OSIRIS programme in his role as Co-PI (with Pearse).

## DECLARATION OF INTERESTS

RP has received research grants and/or honoraria from Edwards Lifesciences, Intersurgical and GloaxoSmithkline, and is a member of the editorial board of the British Journal of Anaesthesia.

## FUNDING

The study was funded by the UK National Institute of Health Research [RP-PG-0217-10001]. It forms the first part in the OSIRIS research programme (*Optimising Shared decision-makIng for high RIsk major Surgery*, https://osiris-programme.org/). The funding agreement ensured the authors’ independence in designing the study, interpreting the data, writing, and publishing the report. The views expressed are those of the author(s) and not necessarily those of the NIHR or the Department of Health and Social Care.

## Notes

### Clinical Protocols

https://bmjopen.bmj.com/content/10/5/e033703.abstract

### Author Declarations

The study received ethical approval from South Central Oxford C Research Ethics Committee (19/SC/0043) of the UK Health Research Authority in February 2019

